# Comparing P300 flashing paradigms in online typing with language models

**DOI:** 10.1101/2022.06.24.22276882

**Authors:** Nand Chandravadia, Shrita Pendekanti, Dustin Roberts, Robert Tran, Saarang Panchavati, Corey Arnold, Nader Pouratian, William Speier

**Affiliations:** Deparment of Computer Science, Columbia University, New York, NY, USA; Department of Radiological Sciences, University of California, Los Angeles, Los Angeles, CA, USA; Department of Neurological Surgery, University of Texas, Southwestern, Dallas, TX, USA

## Abstract

The P300 Speller is a brain-computer interface system that allows victims of motor neuron diseases to regain the ability to communicate by typing characters into a computer by thought. Since the system has a relatively slow typing speed, different stimulus presentation paradigms have been proposed designed to allow users to input information faster by reducing the number of required stimuli or increase signal fidelity. This study compares the typing speeds of the Row-Column, Checkerboard, and Combinatorial Paradigms to examine how their performance compares in online and offline settings. When the different flashing patterns were tested in conjunction with other established optimization techniques such as language models and dynamic stopping, they did not make a significant impact on P300 speller performance. This result could indicate that further performance improvements on the system lie beyond optimizing flashing patterns.

## Introduction

Victims of amyotrophic lateral sclerosis (ALS), brain-stem stroke, and other upper motor neuron diseases lack the ability to vocalize their thoughts and emotion. With a sustained loss of speech, their capacity to write, speak, and laugh is irreversibly impacted. However, the advent of augmentative and alternative communication devices (AAC), such as brain-computer interfaces (BCI), have provided a possible avenue to restore their ability to communicate with the external world.

The P300 speller, an electroencephalogram (EEG)-based BCI, translates neural signals recorded from the scalp into speech in the form of virtual commands on a computer screen [1]. This system utilizes the P300 signal, an endogenous event-related potential (ERP) with a characteristic positive potential after a 300 millisecond delay from stimulus presentation [2]. First introduced by Farwell and Donchin, this system has users attend to a 6×6 matrix composed of alphanumeric characters. The user attends to a character on the matrix while the rows and columns of the matrix flash randomly. Because the target character flashes relatively infrequently in a stream of non-target, repeated stimuli, attending to the target character on the matrix elicits the P300 signal, according the “oddball” paradigm. The P300 signal, observed in the EEG, is then used in classification to detect which character on the matrix was selected. Though the P300 signal is robust, these systems generally have a relatively slow typing speed. Therefore many studies have focused on system optimization, attempting to improve overall system speed.

System optimization studies have traditionally focused on enhancing specific components of the P300 speller apparatus. For instance, Allison et al. modified the matrix size, demonstrating that increasing the size of matrix improves the amplitude of the P300 signal [3]. Lu et al. evaluated the inter-stimulus-interval (ISI), suggesting a longer ISI translates to a both a higher online accuracy and higher characters per minute (CPM) rate [4]. Both Townsend et al. and Jin et al. developed novel flashing patterns, demonstrating significant improvements in bit rate and practical bit rate compared to the traditional row column paradigm (RCP) [5], [6]. Recently, work has shown that a viable strategy of enhancing system performance is to simultaneously combine distinct optimization techniques into a singular method. For instance, Speier et al. tested the performance of a ‘famous faces’ stimulus paradigm integrated with a previously published particle filtering algorithm into a singular approach, establishing that the concatenation of two distinct methodologies into one offers superior results versus both approaches alone [7], [8].

This study surveys the differences in system performance between three proposed flashing patterns: Row-Column Paradigm (RCP), Checkerboard Paradigm (CBP), and the Combinatorial Paradigm (COMB) along with the integration of a language model using a particle filtering algorithm [8]. We hypothesize that the improvements offered by different flashing patterns are negligible in comparison to those from the incorporation of a language model, and therefore that improvements to BCI performance lie outside of flashing pattern optimization.

### Checkerboard Paradigm

The checkerboard paradigm, CBP, was introduced as a way of improving upon the errors associated with the RCP, while concurrently improving overall BCI performance [5]. The goal with the CPB was therefore to design a novel flashing pattern that addressed the constraints associated with the RCP: the adjacency effect and the double flash pattern [9], [10]. The adjacency effect describes situations where flashes of an adjacent row or column (i.e., non-target characters) draws the user’s attention, leading to false-positive P300 signals and ultimately erroneous detections of the intended character [9]. Further, the double flash pattern highlights an inadvertent conundrum associated with the RCP: random sequential row (column) or column (row) flashes can decrease the temporal resolution of the P300 signal [10]. First, because a requisite of the oddball paradigm is the presentation of “deviant stimuli” (i.e., random stimuli), consecutive flashes can impair the detection of the second flash. That is, only the first flash of the target row (column) flash will elicit the P300 signal; the second will not. Kanwisher reported this observation as the repetition blindness phenomenon [11]. In a standard rapid serial presentation (RSVP) task, consecutive stimuli presented with a temporal resolution of less than 500 milliseconds abate the recognition of the succeeding stimuli. In the P300 speller, the flash duration for a single target selection and the ISI are both 62.5 milliseconds, meaning the second flash occurs 125 milliseconds from the onset of the preceding flash, thereby diminishing the ability of the user to resolve the detection of the second flash. Hence, the aim of the CBP sought to mitigate these issues by addressing them in the stimulus design.

The CBP superimposes an imaginary checkerboard over the matrix in such a way that each adjacent character belongs to a different class [5]. Because a checkerboard inherently has an alternating pattern of two colors, the adjacent characters are grouped into two distinct classes. The characters of these two classes then randomly populate one of two corresponding virtual matrices, which the user never observes. These virtual matrices determine the stimulus pattern for each trial (i.e., each target selection). During each target selection, the rows within both virtual matrices are flashed followed by the columns of both virtual matrices. As the rows and columns of the virtual matrices are flashed, the corresponding characters on the real matrix are presented to the user. This methodology reduces the adjacency effect, ensuring that adjacent characters never experience simultaneous flashes, and further safeguards against sequential flashes (i.e., double flashes). However, it requires a larger number of flashes in order to distinguish between each of the characters in the grid.

### Combinatorial Paradigm

The Combinatorial Paradigm (COMB) proposed by Jin et al. utilizes mathematical combinations to minimize the number of flashes per trial with the intention of optimizing the practical bit rate of the system [6]. Reducing the number of flashes per trial would hypothetically improve the selection rate (i.e., due to a reduced number of flashes for classification), leading to an improved PBR (practical bit rate), while still maintaining the vitality of the P300 amplitude. The goal of the COMB paradigm was therefore to optimize the number of target flashes to improve the efficacy of the system.

To choose an optimal number of flashes per trial, Jin et al. used the binomial coefficient of the *x*^*k*^ term of (1 + *x*)^*n*^, where n equals the total number of flashes per trial, and k equals the number of flashes on the target character. Essentially, this translates to:

Since a traditional 6×6 matrix holds up to thirty-six characters, Jin et al. proposed using 7-flash and 9-flash patterns, which locate thirty-five and thirty-six characters, respectively. The 7-flash pattern is based on the combination 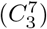, meaning that each trial has seven flashes and the target character flashes three times. Here, a single trial refers to one set of stimuli for a single target selection. Therefore, selecting the character “A” for a single trial should elicit three P300 responses in a set of seven flashes. Since the combination 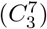 equals thirty-five, the 7-flash pattern locates thirty-five characters in a traditional 6×6 matrix. On the other hand, the 9-flash pattern is based on the combination 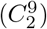, which results in each trial having nine flashes and the target character flashing two times. This combination equals thirty-six, meaning that the corresponding flashing pattern locates thirty-six characters on the traditional 6×6 matrix. The 9-flash pattern locates the same amount of characters as the RCP flash pattern, while the 7-flash pattern locates one less. Likewise, a 12-flash pattern, which mirrors the RCP flash pattern, is modeled as “12 choose 2”, where there are a total of twelve flashes for two target selections per trial – 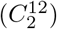. In comparison to the 7- and 9-flash patterns, the 12-flash pattern creates a 71.43% and 33.33% increase in the total number of flashes per trial, respectively.

## Methods

### Subjects

Ten healthy subjects (6 male, 4 female) aged 20 to 35 years old participated in this study. All subjects were cognitively viable with no noticeable neurological deficits. All subjects formally consented to participate. This study was approved by the Institutional Review Board (IRB) at UCLA.

### Data Collection

EEG data were collected from a 32-cap electrode (g.GAMMAcap^2^, Guger Technologies), and signals were amplified with two 16 channel g.tec biosignal amplifiers (Guger Technologies). Signals were sampled at 256 Hz, referenced to the left ear, grounded to AFz, and filtered using a bandpass filter from 0.1 to 60 Hz. BCI2000, a BCI-based development framework, was used for stimulus presentation and data collection [12]. Users were presented with a 6×6 matrix consisting of alphanumeric characters with ‘famous faces’ flashes [13]. Three distinct flashing patterns, RC, CBP, and COMB, were presented to the user to assess divergence in performance (Table 2). Each flash lasted for 62.5 ms with a 62.5 ms ISI, yielding a 125 ms stimulus onset asynchrony (SOA). Subjects completed three training sessions for each flashing pattern—creating a total of nine sessions per subject. For each flashing paradigm, users copied three, ten character words, including spaces. Training data from each session was used for classification in its corresponding flashing pattern. If classification reached a significant benchmark from calibration data, online testing was performed for the trained flashing pattern. In this case, classification was appreciable for all subjects, so all subjects performed three online testing sessions. The order of online testing for each flashing pattern was randomized to dilute the effects of non-familiarity. Classification was performed using a previously established particle filtering (PF) algorithm [8].

### Language Model

In this study, we use a probabilistic automata model as described by Speier et al. [8]. The model employs a directed graph that has states for each substring that starts a word in the corpus, beginning with a blank root node. (Figure 2). Each node has directed edges to nodes that add a single character to the string. For example, if the model only contained the word “CAR,” it would have four states: the root node representing a blank string, “C,” “CA,” and “CAR.” When the word “CAKE” is added to the model, it shares the root node and the “C” and “CA” states, and adds two additional states: “CAK” and “CAKE.” The state “CA” then links to both the states “CAR” and “CAK.” If a state represents a completed word, it will have a link back to the root node to begin a new word. The state “CAR,” for instance, links to the root because “CAR” is a complete word, but it also is the beginning of other words so it has additional links to other states such as “CARD” or “CART.” The relative frequencies of substrings in the Brown English language corpus determined transition probabilities between nodes [28]. For instance, the probability of typing the letter “R” after “CA” has already been entered is determined by dividing the number of occurrences of words that begin with “CAR” by the number of times words start with “CA” in the corpus. Similarly, the probability that a word ends and the state transitions back to the root is the ratio of the number of times that word occurs in the corpus over the number of word occurrences starting with that substring.

### Classifier

Since it is impractical to compute the probability distribution over all possible strings typed by the user in real time, the probability distribution is estimated using the PF classifier. This classifier estimates the probability distribution over possible outputs by sampling a batch of possible realizations of the model (i.e., a batch of output strings that could have been typed by the user). Each of these realizations is called a particle, which contains a pointer to a node in the model and represents one possible configuration of the model at a given time. Each of these particles moves through the language model independently, based on the model’s transition probabilities. Low probability realizations are periodically replaced by more likely realizations by resampling the particles based on weights derived from the observed EEG responses. The algorithm estimates the probability distribution of the possible output strings by finding the proportion of the particles that point to each state after they have moved through the model. In order to determine the probability that the user is attempting to type a given character *x*_*t*_ based on the observed signals, stepwise linear discriminant analysis (SWLDA) is used to select a set of signal features to include in a discriminant function [29]. During training, the algorithm uses ordinary least-squares regression to predict class labels and iteratively adds the most significant features and removes the least significant features until either the target number of features was met or it reached a state where no features were added or removed [10]. The score for flash in for character t, 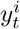, can then be computed as the dot product of the feature weight vector with the features from that trial’s signal. It has been shown that scores can be approximated as independent samples from a Gaussian distribution given the target character [19].

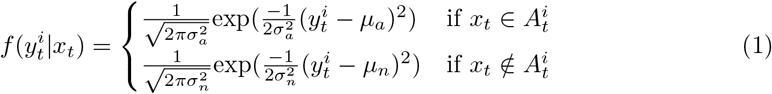

where *μ*_*a*_, 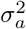, *μ*_*n*_, and 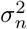 are the means and variances of the distributions for the attended and non-attended flashes, respectively, and 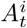 is the set of characters highlighted in flash i. The conditional probability of a target at time t given the EEG signal and the previous target characters *x*_0:*t−*1_ can then be found:

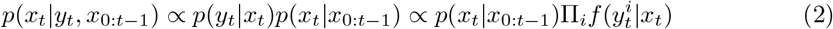

where *p*(*x*_*n*_|*x*_0:*n−*1)_) is the prior probability of character *x*_*n*_ given the previously selected characters, determined from the language model. Because the previous target characters are unknown, it is necessary to compute the probability over all possible output strings. This computation is impractical, so the distribution needs to be estimated using sampling methods such as particle filtering. In particle filtering, a set number of samples (i.e., particles) are created to estimate the distribution over the language model. Each particle j consists of a link to a state in the language model, 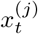; a string consisting of the particle’s state history, 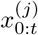, the index of the last time the particle was in the root node, m; and a weight, *w*^(*j*)^. When the system begins, a set of P particles is generated and each is associated with the root node with an empty history and a weight equal to 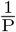. At the start of a new character, a sample character 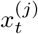 is drawn for each particle from the proposal distribution defined by the language model’s transition probabilities from the particle’s history, 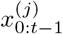.

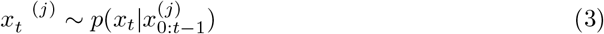

where 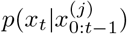 is provided by the language model as in eqn 1. When a particle transitions between states, its pointer changes from the previous state in the model, *x*_(*t−*1)_, to the new state *x*_*t*_. The history for each particle, 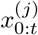, is stored to represent the output character sequence associated with that particle. After each stimulus response, the score for that response, 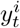, is computed and the probability weight is updated for each of the particles:

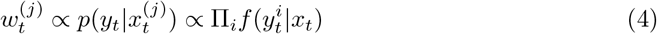

Where 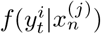 is computed as in Eqn 2. The weights are then normalized and the probability of an output string is found by summing the weights of all particles that correspond to that string.

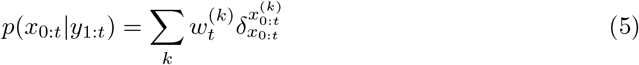

where *δ* is the Kronecker delta. Dynamic classification was implemented by setting a threshold probability, *p*_thresh_, to determine when a decision should be made. The program flashes characters until either the maximum probability exceeds the threshold, or the number of sets of flashes reached the maximum (10 flashes). The classifier then selects the string that satisfied 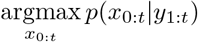. If characters in this output differ from the previous output text, the previous characters are assumed to be errors and are replaced by those in the current string. A new batch of particles, 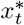, are then sampled from the current particles, *x*_*t*_, based on the weight distribution, *w*_*t*_. Each of the new particles are then assigned an equal weight 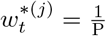. The subject then moves on to the next character and the process then repeats with the new batch of particles. The optimization of *p*_thresh_ is impractical for online experiments, so a previously reported value of 0.95 was used for all trials [16].

### Predictive Spelling

When predictive spelling is added to the model, the same classifier and language model are used, but the projection step is modified in order to estimate the probabilities of potential completed words. When particles are being projected, a proportion, *ρ*, of them continue moving throughout the model until they reach the root node. Note that because particles can move multiple steps in one transition, the length of the particle history can now be greater than t, so it is denoted *n*. Each particle can have different values of *n* and *m*; the subscript *j* is omitted for these values here for simplicity. After projection, the probability distribution over words is found by summing the weights of particles that have been projected forward to completed words

The top *k* of these words are then added to designated locations in the character grid (Figure 1). EEG responses associated with flashing those cells are applied to the particles that have been projected to those words. Particles that were projected to lower probability words are given zero probability and will be replaced during the next resample phase. In this study, the probability of a complete word selection was set empirically to 0.40 and six word suggestions were presented to the user.

**Fig 1.**
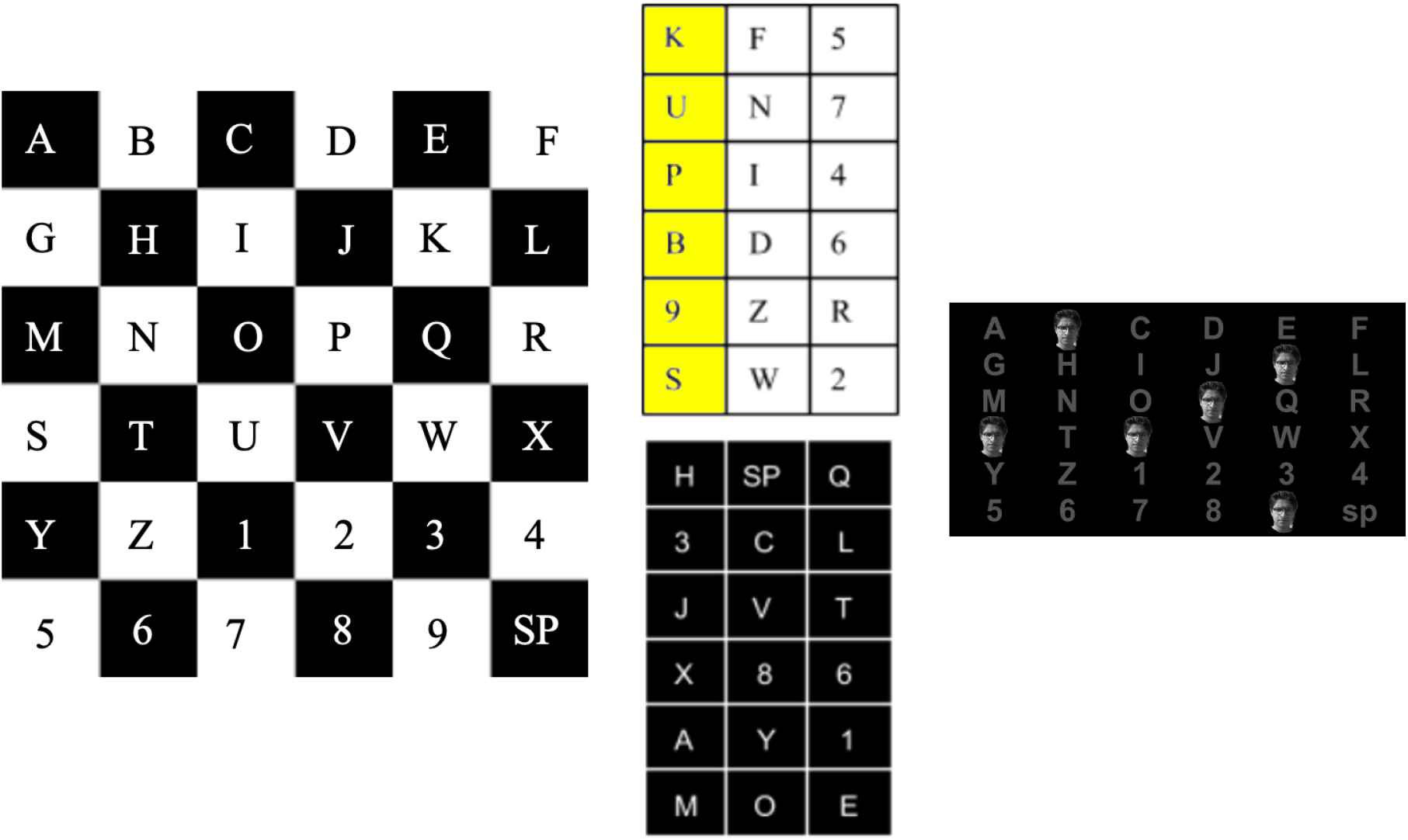
Checkerboard paradigm (CBP) in a 6×6 matrix. **Left:** Matrix with imaginary checkerboard superimposed over it; adjacent characters are assigned to different classes. *Center:* Characters arranged in a virtual matrix, each matrix represents a different class. In this example, the first column has been selected from the top matrix. *Right:* Actual display seen by user; a face is flashed on top of the selected characters from the virtual matrices.

**Fig 2.**
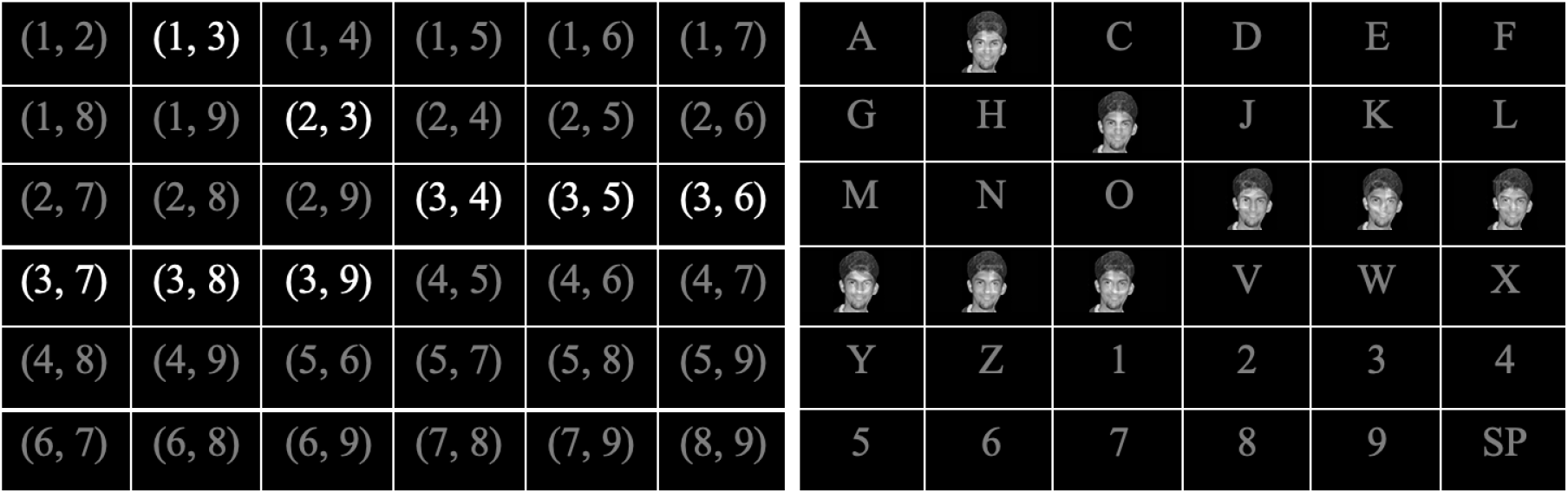
Combinatorial paradigm (COMB) with 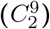 flashing pattern. *Left:* Each character is assigned a unique, two number identifier corresponding to the time it will be flashed. For example (1,3) indicates that the character will be flashed in the first and third flash. For simplicity, the characters in this figure are assigned indices sequentially, in practice the assignment would be random. In this case, we depict the third flash; so all characters corresponding to the number 3 are flashed. *Right:* The output seen by a user; a face is flashed over characters that are assigned flash index 3.

**Fig 3.**
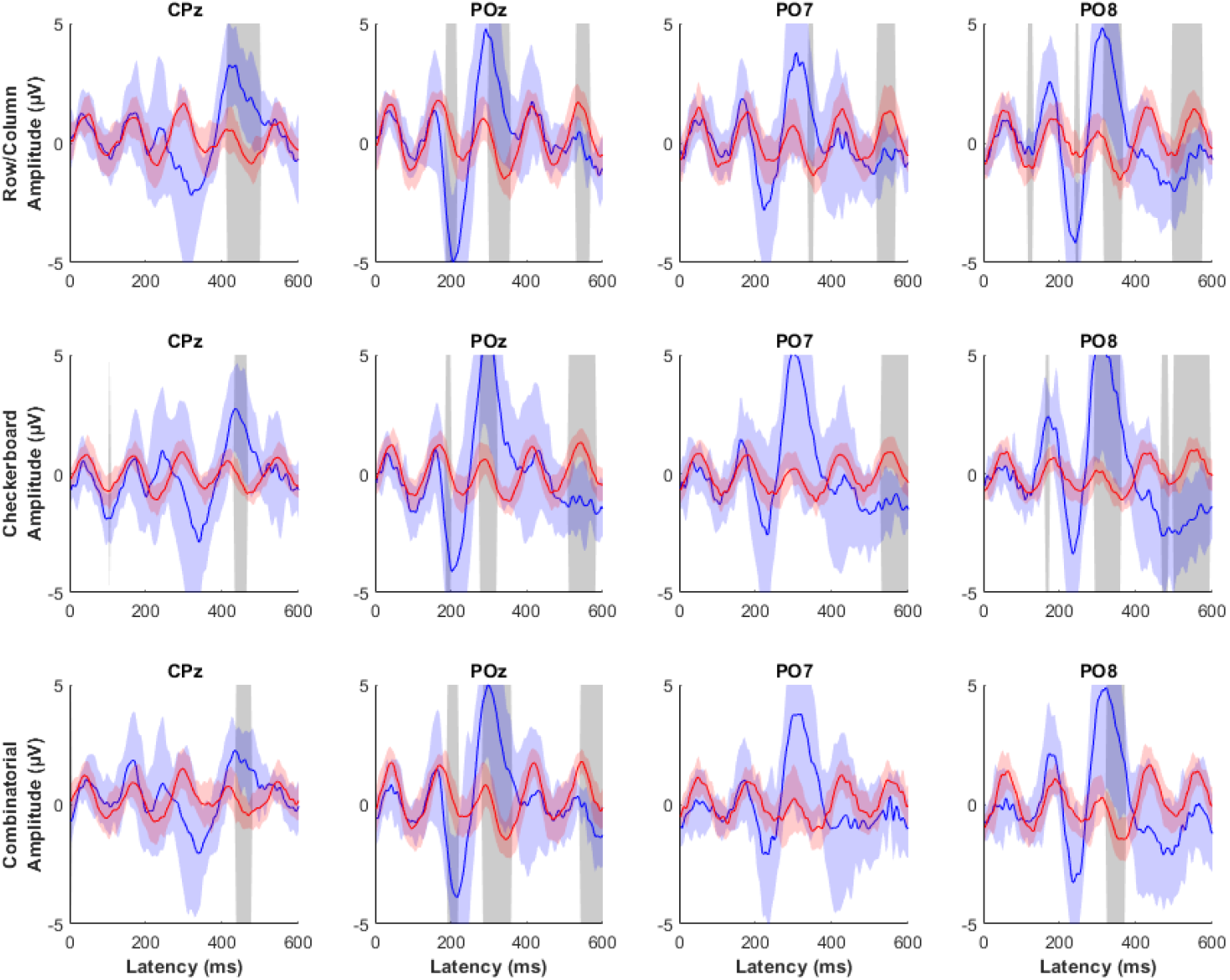
Target P300 waveforms. The average target response for each flashing pattern at CPz, POz, PO7, and PO8 for subject 2 when using the row/column (blue), checkerboard (green), and combinatorial (red) flashing paradigms.

### Evaluation

Evaluation of a BCI system must take into account two factors: the ability of the system to achieve the desired result and the amount of time required to reach that result. Because there is a trade-off between speed and accuracy, evaluation in BCI communication literature is traditionally based on the mutual information between the selected character, x, and the target character, z, referred to as the bit rate (BR).

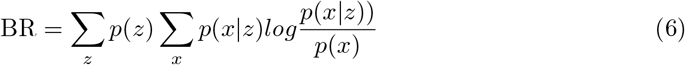

In the most common metric, information transfer rate (ITR), the probabilities for all characters are assumed to be the same 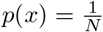 (where N is 36, the size of the alphabet) and errors are assumed to be uniform across all possible characters, so

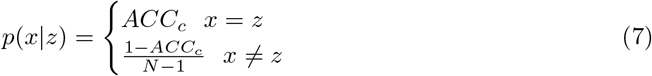

where 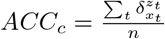 is the single character accuracy and n is the total number of characters selected. This reduces the bit rate to

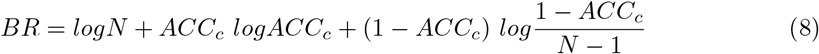

This is then multiplied by the average number of characters selected per minute (CPM=n/time) to produce the ITR [26].

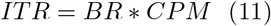

One problem introduced by including PS is that sentences including erroneous word completions could be a different length from the target. Comparing at the character level no longer works in this case. One solution is to base accuracy on Levenshtein distance (LD) (i.e., the minimum number of insertions, deletions, and replacements required to convert x into z) [30]. We then have 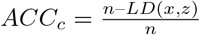 and the equations above hold. It has previously been pointed out that ITR overestimates the amount of information conveyed by the system because characters do not occur with equal frequency [31]. Also, the amount of information that ITR assigns to a word is based largely on the word’s length. This metric assigns a significantly higher amount of information to incorrect strings that are share characters to the target, regardless of whether they make syntactic sense or possibly confuse the meaning (Table 1). An alternative would be to base the metric on word frequency 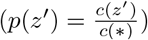. The accuracy can then be computed as the fraction of correct words 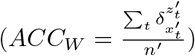, resulting in a conditional probability of a selection:

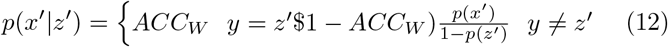

The bit rate then becomes

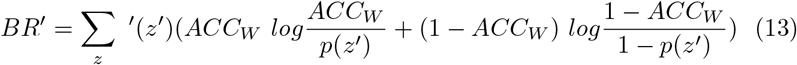

The mutual information can then be found by multiplying by the words selected per 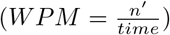.

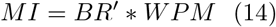

Because the distributions for speeds, accuracies, and bit rates are not normally distributed, significance was tested for all metrics using the nonparametric Kruskal-Wallis test.

**Table 1.** The offline selection rate (SR), accuracy (ACC), and information transfer rate (ITR), for each flashing pattern.

| Subjects | SR (Characters/minute) |  |  | ACC (%) |  |  | ITR (bits/minute) |  |  |
| --- | --- | --- | --- | --- | --- | --- | --- | --- | --- |
|  | RCP | CBP | COMB | RCP | CBP | COMB | RCP | CBP | COMB |
| 1 | 13.46 | 13.87 | 13.91 | 100.00 | 95 | 100.00 | 69.61 | 64.19 | 71.93 |
| 2 | 13.32 | 12.94 | 15.07 | 95 | 100.00 | 100.00 | 61.61 | 66.89 | 77.91 |
| 3 | 14.769 | 13.75 | 15.71 | 100.00 | 100.00 | 95 | 76.35 | 71.10 | 72.70 |
| 4 | 11.12 | 9.80 | 13.79 | 100.00 | 100.00 | 100.00 | 57.51 | 50.64 | 71.31 |
| 5 | 12.938 | 13.52 | 12.97 | 95 | 95 | 100.00 | 59.86 | 62.56 | 67.07 |
| 6 | 14.90 | 14.44 | 14.55 | 95 | 100.00 | 100.00 | 68.97 | 74.63 | 75.20 |
| 7 | 14.747 | 14.24 | 13.87 | 100.00 | 100.00 | 100.00 | 76.24 | 73.63 | 71.72 |
| 8 | 13.278 | 12.70 | 13.93 | 100.00 | 100.00 | 100.00 | 68.64 | 65.65 | 72.03 |
| 9 | 14.307 | 12.57 | 14.14 | 95 | 100.00 | 95 | 66.20 | 64.96 | 65.42 |
| 10 | 12.55 | 14.63 | 13.56 | 100.00 | 95 | 100.00 | 64.87 | 67.71 | 70.10 |
| Mean | 13.54 | 13.25 | 14.15 | 98 | 98.5 | 99 | 66.99 | 66.20 | 71.54 |

**Table 2.** The online selection rate (SR), accuracy (ACC), and information transfer rate (ITR), for each flashing pattern.

| Subjects | SR (Characters/minute) |  |  | ACC (%) |  |  | ITR (bits/minute) |  |  |
| --- | --- | --- | --- | --- | --- | --- | --- | --- | --- |
|  | RCP | CBP | COMB | RCP | CBP | COMB | RCP | CBP | COMB |
| 1 | 14.65 | 15.08 | 15.86 | 100.00 | 100.00 | 97.27 | 75.73 | 77.94 | 76.91 |
| 2 | 16.46 | 16.11 | 17.14 | 100.00 | 100.00 | 100.00 | 85.11 | 83.3 | 88.6 |
| 3 | 16.79 | 15.43 | 16.61 | 100.00 | 100.00 | 100.00 | 86.82 | 79.76 | 85.89 |
| 4 | 12.38 | 11.48 | 10.63 | 98.59 | 100.00 | 94.74 | 61.77 | 59.36 | 48.93 |
| 5 | 15.05 | 13.89 | 12.41 | 81.25 | 100.00 | 82.50 | 52.87 | 71.79 | 44.71 |
| 6 | 15.34 | 13.63 | 14.72 | 100.00 | 100.00 | 100.00 | 79.3 | 70.48 | 76.11 |
| 7 | 15.35 | 13.19 | 12.77 | 100.00 | 100.00 | 98.57 | 79.35 | 68.21 | 63.7 |
| 8 | 13.39 | 12.58 | 14.72 | 95.95 | 100.00 | 76.92 | 63.17 | 65.06 | 47.2 |
| 9 | 13.71 | 14.51 | 14.79 | 100.00 | 100.00 | 90.67 | 70.88 | 75.02 | 62.78 |
| 10 | 12.79 | 12.18 | 10.01 | 100.00 | 100.00 | 97.67 | 66.13 | 62.99 | 48.97 |
| Mean | 14.59 | 13.81 | 13.97 | 97.58 | 100.00 | 93.83 | 72.11 | 71.39 | 64.38 |

## Results

### Offline Analysis

In the offline analysis, no one paradigm significantly outperformed any other across the three measured metrics. Table 1 shows the offline selection rate (in characters per min), accuracy, and ITR for each flashing pattern. The differences in median SR between the three flashing paradigms were not found to be statistically significant (H = 2.96, p = 0.227). The accuracy across the different paradigms, while high, are also not significantly different (H = 0.581, 0.748). While the combinatorial paradigm has a slightly better accuracy, there are no significant differences in ITR between the three paradigms (H = 5.257, p = 0.0722).

### Online Analysis

Table 2 shows the online selection rate (in characters per min), accuracy, and ITR for each flashing pattern. Despite the RCP yielding the highest mean SR, there were no significant differences in the mean SR for any of the flashing patterns (H = 0.98, p = 0.611) (Table 2). In contrast, the difference in the median accuracy across the flashing patterns was found to be statistically significant (H = 7.399, p = .025). Pairwise Mann-Whitney tests between each of the flashing patterns demonstrate that the CBP pattern was significantly higher than the COMB flashing pattern (p = .0045). There are no appreciable differences in accuracy between RCP and CBP across (p = .271) and between RCP and COMB for (p = .112). Further, the mean ITR, which is a function of both accuracy and SR, was not significantly different for any of the flashing patterns (H = 1.46, p = 0.481), consistent with SR and the offline paradigm. Therefore, no appreciable differences were detected in BCI performance among each of the flashing patterns.

### Waveform Analysis

The P300 signal for each flashing pattern was evaluated at CPz, POz, PO7, and PO8 to examine for meaningful differences in the amplitudes of the waveforms [14]. Stimulus responses during online sessions were grouped based on whether the stimulus contained the target character. The average attended and non-attended responses were calculated for each subject and a global average was produced across subjects for each channel. Significance was tested at each latency to determine whether attended and non-attended responses differed significantly using Wilcoxon signed rank tests correcting for multiple comparisons using false discovery rate.

For all three stimulus paradigms, there were significant differences between attended and non-attended stimulus responses. Each of the four channels had a large positive peak preceded by a smaller negative peak in the attended responses. In the parieto-occipital channels, the negative peak occurred at a latency of approximately 200 ms and the positive peak at a latency of 300 ms. In the CPz channel, these peaks were slightly later, occurring at approximately 300 ms and 400 ms latencies, respectively. In the CPz, POz, and PO8 channels, the positive peak was significantly different from the non-attended response. The peak in the PO7 channel was not statistically significant, most likely because of a high variance across subjects.

The average signals were compared across the responses for the three stimulus paradigms. While the positive peaks for the parieto-occipital channels were generally larger for the checkerboard paradigm, no significant trend was seen between the three groups. This result suggests that the stimulus paradigm does not significantly affect the stimulus response produced.

## Discussion

A robust, clinically viable BCI speller requires high accuracy (*>*90%), and speed (at least 15-19 characters per minute) [15]. Although the functional utility of the P300 speller has been demonstrated in invasive conditions, specifically with signals acquired with electrocorticography (ECoG), the long-term safety and utility has yet to be determined. In order to ameliorate the risks of an invasive procedure, several studies aim to optimize the utility of a P300 speller with a non-invasive, EEG-based paradigm. Much work has been done to try and optimize the flashing pattern used, but has yielded mixed results [5], [6].

Our study aimed to provide a meaningful, standardized comparison of performance for each flashing pattern, incorporating optimization methodologies that have been shown to enhance performance. In our study, alternative flashing paradigms did not significantly improve typing performance in a system with dynamic stopping and language model priors. The mean online selection rate, mean online accuracy, and mean online ITR were not significantly different for any of the three flashing patterns. This observation contrasts with reports from both Townsend (2010) and Jin (2010) that the traditional RCP flashing pattern failed to meet equivalent performance standards compared to the CBP and COMB flashing patterns, respectively [5], [6].

Townsend (2010) reported that the CBP flashing pattern yielded both a greater online accuracy and practical bit rate [5]. In a 72 character grid, there are 24 flashes per target selection in the CBP flash pattern compared 17 flashes per target selection the RCP flash pattern. Because the number of flashes in the CBP is higher than in the RCP, this would naturally lead to a greater time duration for each target selection, leading to lower SR. Leveraging dynamic stopping, where the number of flashes per target selection is modulated by the classification threshold, would dilute this disparity, normalizing the selection rate for the CBP and RCP flashing patterns. While we find that the CBP pattern has significantly higher accuracy, we hypothesize that the excellent accuracy performance of the CBP pattern in the online paradigm is due to the fact that CBP optimizes for accuracy while making significant concessions to speed.

Jin (2010) stated that mean offline practical bit rate was significantly different for the 9-flash pattern compared to the 12-flash pattern (RCP), as a result of the diminished number of flashes required for a character selection – a 33.33% decrease in the number of flashes per selection [6]. Although there were fewer characters required for each character selection, this did not necessarily translate to a higher online selection rate.

Our results suggest that dynamic stopping, where the number of flashes per target selection changes as a function of a classification threshold needed to select a character, reduces the performance effects of a nine-flash pattern with static number of flashes. Dynamic stopping allows the system make decisions without needing to wait for a required number of flashes, thereby reducing the impact of the flashing pattern on performance.

## Conclusion

This study shows that when used in conjunction with other established methods, proposed flashing paradigms do not make a significant impact on P300 speller performance. A large contributing factor to this phenomenon could be that dynamic stopping allows the system to make decisions without needing to wait for a required number of flashes, reducing the impact of the flashing paradigm. This result likely implies that current bottlenecks in P300 speller performance lie outside the type of flashing paradigm used, and that optimization methods should be focused on improvements to language models and predictive spelling.

## Data Availability

All data produced in the present study are available upon reasonable request to the authors.

## Acknowledgments

This material is based upon work supported by the National Science Foundation Graduate Research Fellowship Program under Grant No. (DGE-2034835). Any opinions, findings, and conclusions or recommendations expressed in this material are those of the author(s) and do not necessarily reflect the views of the National Science Foundation.

